# Out-of-pocket prescription medicine expenditure amongst community-dwelling adults: findings from The Irish Longitudinal Study on Ageing (TILDA) in 2016

**DOI:** 10.1101/2024.07.11.24310220

**Authors:** James Larkin, Ciaran Prendergast, Logan T. Murry, Michelle Flood, Barbara Clyne, Sara Burke, Conor Keegan, Fiona Boland, Tom Fahey, Nav Persaud, Rose Anne Kenny, Frank Moriarty

## Abstract

**Background:** The number of prescription medicines prescribed to older adults is increasing in Ireland and other countries. This is leading to higher out-of-pocket prescription medicine expenditure for older adults, which has several negative consequences including cost-related non-adherence. This study aimed to characterise out-of-pocket prescription medicine payments, and examine their relationship with entitlements, multimorbidity and adherence.

**Methods:** This cross-sectional study used 2016 data from a nationally-representative sample of adults in Ireland aged ≥50 years. Descriptive statistics and regression models were used to describe out-of-pocket prescription medicine payments and assess the association between out-of-pocket prescription medicine payments and the following variables: healthcare entitlements, multimorbidity, and cost-related non-adherence.

**Results:** There were 5,668 eligible participants. Median annual out-of-pocket prescription medicine expenditure was €144 (IQR: €0-€312). A generalised linear model showed that, amongst those with out-of-pocket prescription medicine expenditure, having fewer healthcare entitlements was associated with 4.74 (95% CI: 4.37-5.15) times higher out-of-pocket prescription medicine expenditure. Overall, 1.7% (n=89) of participants reported cost-related non-adherence in the previous year. A multivariable model found no significant associations between any variables and cost-related non-adherence.

**Conclusions:** Those with entitlements to subsidised prescription medicines had much lower out-of-pocket prescription medicine expenditure. This highlights the benefits of expanding healthcare entitlements and ensuring uptake of entitlements by those with eligibility.

## 1. Background

In most OECD countries, including Ireland, per capita pharmaceutical expenditure increased between 2010 and 2020,^1^ with these trends expected to continue.^2^ The primary drivers of increased per capita pharmaceutical expenditure are the development of expensive drugs^3^ and the increased use of prescription medicines.^4^ In Ireland, prescription medicine use has greatly increased in recent decades. For example, the concurrent use of ≥5 medications increased amongst those aged ≥65 years from an estimated 18% of the population to 60% between 1997 and 2012.^5^ These rates have further increased in recent years in Ireland^6^ and in other countries.^7^ Likely reasons include populations ageing^8^ and the increasing prevalence of multimorbidity (≥2e chronic conditions in an individual).^9-11^ Despite theis, multimorbidity is an often-overlooked complicating factor in medication regimens^12^ and in eligibility to publicly-funded schemes.^13,14^

These complexities can contribute to varied approaches to how health systems provide and finance medicines for their population, which can include public financing, health insurance and user fees.^15^ Medications are among the most expensive components of healthcare when considering out-of-pocket payments.^16-18^ In Ireland, which has a mixed public-private healthcare system (Table 1), medicines have consistently accounted for the highest share of out-of-pocket healthcare spending for the poorest quintiles.^19^ Between 2009 and 2016 there was a large increase in out-of-pocket healthcare payments in Ireland, a signification proportion of this increase attributed to prescription medicines.^20^

**Table 1.** Details of prescription medicine entitlement schemes.

|  | **DPS<sup>25</sup>** | **GMS Scheme<sup>26</sup>** | **HTD Scheme<sup>27</sup>** | **HAA<sup>28</sup>** | **LTI Scheme<sup>29</sup>** |
| --- | --- | --- | --- | --- | --- |
| **Applicable medicines** | All | All | ‘High Tech’ Drugs | All | medicines to treat one of 16 specified health conditions |
| **Eligibility** | Those without GMS entitlements | Means tested based on income, age and household structure | Prescribed ‘High Tech’ Drugs by a hospital consultant | Those affected by contaminated blood products provided by the State | Individuals with any one of 16 specified health conditions |
| **Monthly limit** | €144 per household per month | €25 per household per month | €144 for DPS, €0 for all other groups | €0 | €0 for applicable medicines |
| **Co-payment limit** | N.A. | €2.50 per medicine dispensed | N.A. | €0 | €0 for applicable medicines |

DPS=Drug Payments Scheme GMS= General Medical Services HTD=High Tech Drugs HAA=Health Amendment Act
LTI=Long Term Illness

High out-of-pocket payments can have a range of consequences. A review of the Irish health system concluded that the burden of out-of-pocket payments for healthcare was *catastrophic* (total annual out-of-pocket health payments exceeding 40% of a household’s non-subsistence income) for 1.2% of households, while a further 1% were pushed into poverty by out-of-pocket healthcare payments.^19^ Out-of-pocket payments can also cause cost-related non-adherence/attendance, i.e. not accessing or using recommended healthcare services/interventions due to cost, which can ultimately increase healthcare costs due to negative health outcomes.^21^ A review of studies in eleven high-income countries found that prevalence of cost-related non-adherence to prescribed medicines ranged from 1.6-16.8%, with a higher prevalence among lower incomes populations and lower prevalence amongst older adults.^22^ A study examining the introduction of small prescription medicine co-payments in Ireland found that they reduced adherence by between 2-10% depending on the medication.^23^

### 1.1 Healthcare Coverage in Ireland

Healthcare entitlements in Ireland are ‘extremely complex’.^19^ Prescription medicines are mainly funded through public healthcare entitlements and out-of-pocket payments.^19^ In 2016, 36% of the population had General Medical Services (GMS) scheme entitlements and these were primarily low-income groups.^24^ The GMS scheme provides for access to prescription medicines for a low co-payment (full details in Table 1 and eBox 1). Those without GMS entitlements are eligible for the Drugs Payment Scheme (DPS), which, in 2016, meant a monthly prescription medicine payment cap of €144 per household. Details of other relevant schemes are in Table 1 and eBox 1.

### 1.2 Aim

Affordability and multimorbidity are large drivers of patients choosing to reduce, delay, or cease their prescribed medication regimen.^30,31^ However, to our knowledge a detailed analysis of out-of-pocket medicine expenditure and the divergent causes of cost-related non-adherence have not been modelled in the context of Ireland’s complex healthcare entitlements system. This study aimed to characterise out-of-pocket payments for prescription medicines, and examine their relationship with entitlements, multimorbidity and adherence.

## 2. Methods

### 2.1 Study design and participants

This is a cross-sectional study, reported according to the Strengthening the Reporting of Observational Studies in Epidemiology (STROBE) guidelines.^32^ It uses data from wave 4 of The Irish Longitudinal Study on Ageing (TILDA),^33^ a nationally-representative cohort study of adults in Ireland aged ≥50 years. Wave 4 data collection took place in 2016 using computer aided personal interviewing (CAPI). CAPI documentation is available on the TILDA website.^34^

### 2.2 Participants

Participants recruited at baseline were aged ≥50 years, as well as their spouses or partners of any age. Households were sampled based on a random selection process using a national geodirectory of residential addresses. Participants in residential care settings were excluded from analysis due to the likely difference between their healthcare utilisation patterns and the patterns of community-dwelling adults. Participants were also excluded if they had not answered the question on out-of-pocket prescription medicine spending or stated they do not know their expenditure, or if there had a significant cognitive impairment and were therefore not asked this question.

### 2.3 Variables and data sources

#### 2.3.1 Outcomes

The primary outcome variable, out-of-pocket prescription medicine expenditure was ascertained during the CAPI where participants were asked “*Not counting health insurance refunds, on average about how much do you pay out-of-pocket for your prescribed drugs per month*?” This amount was multiplied by 12 to estimate annual expenditure. Full details of medicines questions are in Appendix A, eBox 2. With regard to outliers, data (N=13) was removed if a participant’s monthly prescription medicine expenditure was above €288 (twice the Drugs Payment Scheme limit of €144), unless there was other information provided by the participant to suggest it was plausible or an error had been made (e.g. a decimal point error in recording verbally-reported expenditure). Example cases are provided in Appendix A, eBox 3.

Participants were also asked about cost-related non-adherence: “*In the last 12 months, have you ever received a prescription from your GP that you didn’t fill with the pharmacy because you thought that the medication was too expensive*?” The response options were yes, no or don’t know/refused.

#### 2.3.2 Exposure

The two primary exposures were healthcare entitlements and multimorbidity. Healthcare entitlements were ascertained by asking about the GMS scheme, the GP visit card, the Health Amendment Act, the LTI scheme and private health insurance (questions details in Appendix A, eBox 4). Details of entitlements are in Table 1 and eBox 1. For analysis, those covered under the Health Amendment Act were grouped with the GMS scheme. Entitlement was categorised as GMS scheme, GP visit card, or neither of these (as mutually exclusive groups), and additionally, presence of LTI scheme entitlement and/or private health insurance. *GP Visit Cards* entitle holders to free access to general practitioners (GPs) but provides no specific medication cover.^35^ There is limited clarity onmedication subsidies provided by private health insurance^36,37^ though they are likely to be limited as most insurance plans are hospital plans.^38^

The number and type of health conditions was ascertained by asking participants to report doctor-diagnosed conditions (Appendix A, eBox 5). The condition list for analysis was developed by combining some of the 36 conditions asked about to give 21 broader conditions (Appendix A, eBox 6) in line with previous TILDA research.^39^ For regression analyses, the number of chronic conditions was included as a single count variable. For descriptive analysis, number of chronic conditions was analysed as a count variable and was also grouped into categories (0, 1, 2, 3+ conditions). A binary variable for presence of complex multimorbidity was also included. Complex multimorbidity was defined as the presence of at least three chronic conditions in an individual with a minimum of three conditions each primarily affecting one distinct body system (defined by the World Health Organization’s International Classification of Primary Care-2, see Appendix A, eBox 7).^40^

#### 2.2.3 Covariates

Participants also reported ‘regular’ medications that they take ‘every day or every week’. These responses were summarised to derive the number of regular prescribed medications likely to incur expenditure.

There were several demographic questions including sex, age, urban/rural residence and marital status. Participants also provided details of overall household income. Equivalised household income was used for descriptive analysis, which is an adjusted measure based on the OECD-modified equivalence scale for household size:^41^ household income is divided by number of people in the household, where a weight of 1 is applied to the first adult, 0.5 for each additional adult and 0.3 for each child. Level of educational attainment was also used as a proxy for socioeconomic status. For the residence variable this was operationalised using two categories: urban and not urban.

Total out-of-pocket health expenditure was determined as an individual’s total reported expenditure on prescribed medications, GP visits, emergency department care, specialist medical consultations, and hospital outpatients and inpatient care reported over 12 months preceding their interview (question details in Appendix A, eBoxes 2 and 8).

### 2.4 Statistical analysis

Firstly, descriptive statistics were generated to describe variables of interest. This included a breakdown for each exposure variable within each age-bracket, sex, education level, area of residence (urban/non-urban), marital status, and number of regular medications (grouped in quintiles). Mean and median out-of-pocket prescription medicine expenditure are also provided for the above variables and each exposure variable.

Descriptive statistics were also generated for any out-of-pocket prescription medicine expenditure, out-of-pocket prescription medicine expenditure as a proportion of all out-of-pocket health expenditure, and financial burden of prescription medicines. Financial burden of prescription medicines is the percentage of equivalised household income spent on prescription medicines (winsorised at 100%); previously it has been applied to out-of-pocket healthcare expenditure overall.^17,18^ Equivalent descriptive statistics were generated for participants reporting cost-related non-adherence in the previous 12 months.

A two-part regression model was used to analyse the independent association of each category of healthcare entitlement and number of chronic conditions with out-of-pocket prescription medicine expenditure. The first part was a logit regression to model the binary measure of whether a person had any out-of-pocket prescription medicine expenditure or not. The second part was a generalised linear model (GLM) with log-link and gamma distributed errors which models out-of-pocket prescription medicine expenditure amongst those with any out-of-pocket prescription medicine expenditure. This two-part model allows for the presence of a large number of cases with the value zero, which often occurs for expenditure data.^42^ The two models adjusted for covariates including sex, age, education level, number of prescribed medicines, urban residence and marital status. A sensitivity analysis of the two models was conducted with number of chronic conditions included as a categorical variable.

A logit regression was conducted modelling the binary outcome of cost-related non-adherence. The primary independent variable was out-of-pocket prescription medicine expenditure, parameterised as any expenditure, and amount of expenditure. The model controlled for non-medication out-of-pocket healthcare expenditure (parameterised as any expenditure and the amount of expenditure), healthcare entitlements, demographics, and number of prescribed medications. Complete-case analysis was used for all regression models. An exploratory analysis was conducted which involved univariate logit models with the same independent variables and cost-related non-adherence as the dependent variable.

## 3. Results

There were 5,668 eligible participants, after exclusion of participants in residential care settings (n=78) and those who did not provide details of out-of-pocket prescription medicine expenditure (n=153). The mean age of participants was 68.1 years (SD=8.9), 55.6% (n=3,153) were female and 46.5% (n=2,632) had GMS entitlements. Overall, 30.6% (n=1,734) were taking ≥4 regular medications and the mean number of chronic conditions was 2.1 (SD=1.6).

Median annual out-of-pocket prescription medicine expenditure was €144 (IQR: €0-€312). Median annual out-of-pocket prescription medicine expenditure for those with GMS entitlement was €120 (IQR: €60-€240) compared to €480 (IQR: €180-€1200) for those with a GP visit card and €168 (IQR: €0-€600) for those with eligibility for neither scheme. With regard to multimorbidity, median annual out-of-pocket prescription medicine expenditure ranged from €0 (IQR: €0-€72) for those with no conditions to €240 (IQR: €120-€456) for those with three or more chronic conditions, and this pattern persisted across healthcare entitlement categories (Figure 1). Median annual out-of-pocket prescription medicine expenditure for those on six or more regular medicines was €300 (IQR: €204-€720). Full demographic and out-of-pocket prescription medicine expenditure details are in Table 2. Prevalence of individual chronic conditions and summaries of out-of-pocket prescription medicine expenditure is in Appendix A, eTable 1.

**Figure 1.**
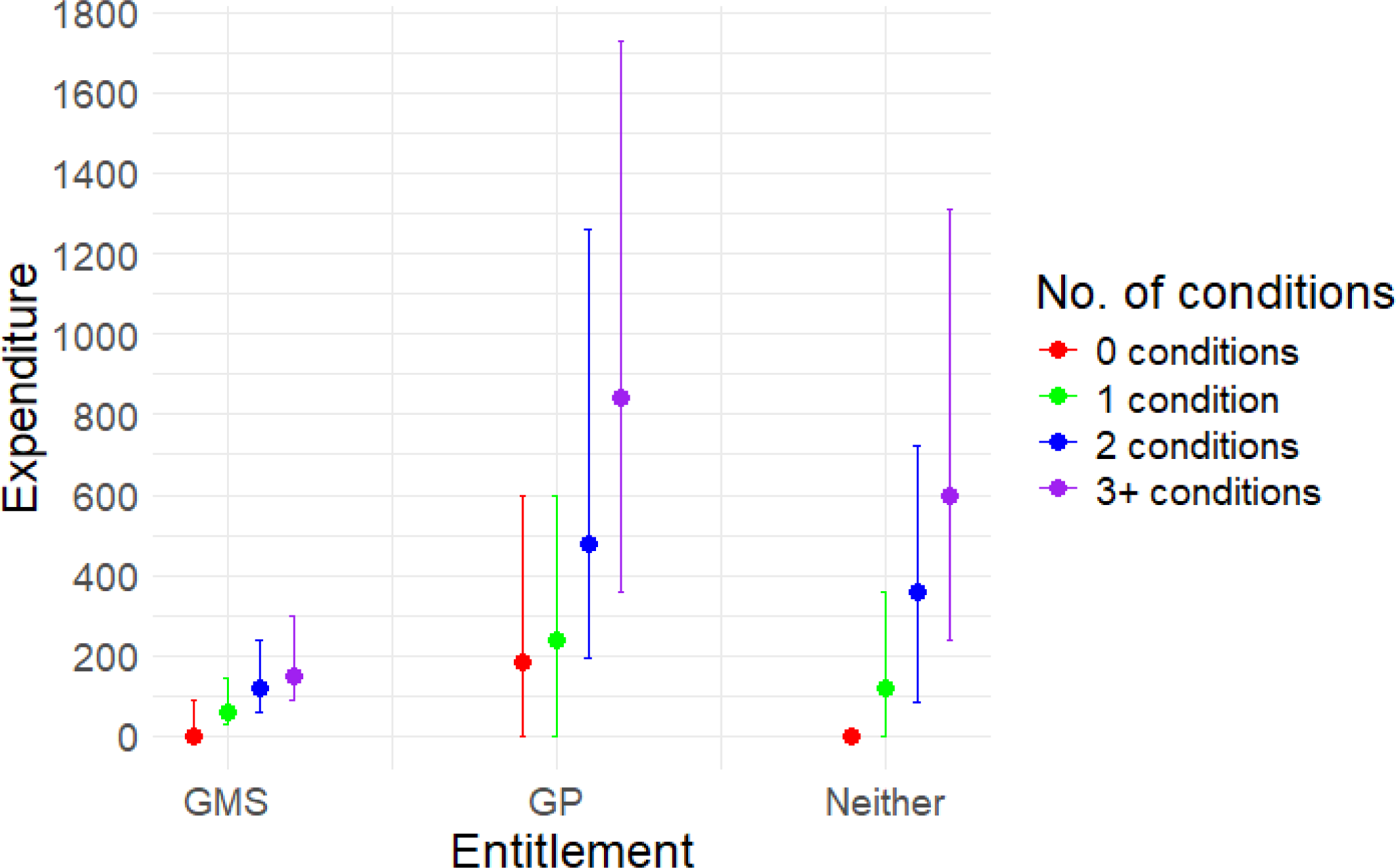
Median (interquartile ranges) out-of-pocket prescription medicine expenditure across healthcare entitlement categories.

**Table 2.** Demographic and entitlement characteristics of sample broken down by out-of-pocket (OOP) prescription medicine expenditure.

|  | Total % (n) | Mean OOP Prescription Medicine Expenditure (SD) | Median OOP Prescription Medicine Expenditure (IQR) |
| --- | --- | --- | --- |
| Age (years) |  |  |  |
| <60 | 19.0 (1,075) | €254 (€439) | €60 (€0-300) |
| 60-69 | 40.9 (2,318) | €330 (€486) | €139.2 (€0-360) |
| 70-79 | 27.8 (1,574) | €325 (€464) | €153 (€60-300) |
| 80-89 | 11.2 (637) | €378 (€500) | €204 (€120-300) |
| 90+ | 1.1 (64) | €246 (€335) | €147 (€90-300) |
| Sex |  |  |  |
| Female | 55.6 (3,153) | €310 (€459) | €144 (€0-300) |
| Male | 44.4 (2,515) | €329 (€489) | €144 (€0-360) |
| Education |  |  |  |
| Primary/none | 23.6 (1,332) | €241 (€343) | €150 (€60-300) |
| Secondary | 39.6 (2,240) | €306 (€469) | €120 (€0-300) |
| Third/higher | 36.9 (2,096) | €381 (€535) | €156 (€0-480) |
| Area of residence |  |  |  |
| Urban | 50.7 (2,873) | €366 (€518) | €168 (€0-360) |
| Not Urban* | 49.3 (2,795) | €270 (€415) | €120 (€0-300) |
| Marital Status |  |  |  |
| Partnered | 69.0 (3,911) | €341 (€504) | €144 (€0-360) |
| Not Partnered | 31.0 (1,757) | €269 (€389) | €150 (€30-300) |
| Private Health Insurance |  |  |  |
| Yes | 60.4 (3,425) | €401 (€540) | €180 (€0-480) |
| No | 39.6 (2,242) | €193 (€304) | €120 (€24-240) |
| Long Term Illness (LTI) Scheme |  |  |  |
| Yes | 7.1 (404) | €307 (€489) | €120 (€12-300) |
| No | 92.9 (5,264) | €319 (€471) | €144 (€0-312) |
| GMS & GP Entitlements |  |  |  |
| GMS Scheme | 46.5 (2,632) | €150 (€184) | €120 (€60-240) |
| GP Visit Card | 10.7 (604) | €722 (€652) | €480 (€180-1200) |
| Neither | 42.8 (2,425) | €401 (€549) | €168 (€0-600) |
| Number of Prescription Medicines (quintiles)** |  |  |  |
| 0 regular medicines | 25.2 (1,429) | €36 (€150) | €0 (€0-0) |
| 1 regular medicine | 17.0 (965) | €235 (€362) | €120 (€30-240) |
| 2-3 regular medicines | 27.2 (1,539) | €389 (€457) | €240 (€90-480) |
| 4-5 regular medicines | 16.7 (948) | €514 (€554) | €240 (€144-720) |
| 6+ regular medicines | 13.9 (786) | €562 (€601) | €300 (€204-720) |
| Number of Chronic Conditions |  |  |  |
| 0 chronic conditions | 17.1 (967) | €111 (€300) | €0 (€0-72) |
| 1 chronic condition | 24.9 (1,411) | €251 (€410) | €120 (€0-300) |
| 2 chronic conditions | 23.4 (1,327) | €374 (€489) | €180 (€60-432) |
| 3+ chronic conditions | 34.6 (1,963) | €432 (€528) | €240 (€120-456) |
| Complex Multimorbidity |  |  |  |
| Yes | 2.0 (114) | €360 (€433) | €240 (€120-300) |
| No | 98.0 (5,554) | €318 (€473) | €144 (€0-312) |

\*Not urban included those who described themselves as rural (n=2,648) and who had missing data (n=147) for this variable.
\*\* Excluded medications not covered by state drug schemes as these are primarily over-the-counter medicines and supplements. Also excluded medications always indicated for a condition covered by the LTI scheme.
Note: SD=Standard Deviation. IQR=Interquartile Range. GMS=General Medical Services. GP= General Practitioner.

Figure 1 shows median out-of-pocket prescription medicine expenditure across three healthcare entitlement categories: GMS scheme, GP Visit Card and neither.

The median financial burden (percentage of equivalised household income spent on prescription medicines) was 1.3% (IQR=0.0%-3.5%). Mean medicine spend as a percentage of total OOP healthcare spend was 68.4% (SD=37.8%). Descriptive statistics for financial burden and medicine spend as a percentage of total OOP healthcare spend, broken down by demographic variables, are in Appendix A, eTable 3.

### 3.1 Modelling out-of-pocket prescription medicine expenditure

The analysis showed a statistically significant negative association between being neither GMS eligible nor having a GP visit-card and likelihood of any out-of-pocket prescription medicine expenditure (OR: 0.63, 95%CI: 0.47-0.83), when compared to those with GMS entitlements (Table 3, model a). Compared to GMS eligibility, there was no association between having a GP visit card and the likelihood of any out-of-pocket prescription medicine expenditure (OR: 0.83, 95%CI: 0.56-1.23). There was a statistically significant negative association seen between being eligible for the LTI scheme and reporting any out-of-pocket prescription medicines expenditure (OR: 0.64, 95%CI: 0.44-0.94). Additionally, a statistically significant positive association was found between number of chronic conditions and likelihood of any out-of-pocket prescription medicine expenditure (OR: 1.47, 95%CI: 1.31-1.64).

**Table 3.** Logit and generalised linear models assessing associations with out-of-pocket (OOP) prescription medicine expenditure.

|  | a) Any OOP expenditure |  | b) Value of OOP expenditure<sup>a</sup> |  |
| --- | --- | --- | --- | --- |
|  | Odds ratio (95%CI) | p value | Rate ratio (95%CI) | p value |
| **Healthcare entitlements (Ref: GMS scheme)** |  |  |  |  |
| GP visit card-holder | 0.83 (0.56-1.23) | 0.353 | 4.70 (4.25-5.19) | <0.001 |
| Neither GMS scheme nor GP visit card | 0.63 (0.47-0.83) | 0.001 | 4.74 (4.37-5.15) | <0.001 |
| **LTI scheme<sup>b</sup>** | 0.64 (0.44-0.94) | 0.021 | 0.98 (0.87-1.09) | 0.693 |
| **Private Health Insurance<sup>b</sup>** | 1.11 (0.86-1.42) | 0.436 | 1.07 (1.00-1.15) | 0.056 |
| **Age (years)** | 1.00 (0.99-1.02) | 0.862 | 1.01 (1.00-1.01) | 0.009 |
| **Female sex<sup>b</sup>** | 0.75 (0.61-0.93) | 0.008 | 0.91 (0.86-0.97) | 0.004 |
| **Education (Ref: primary/none)** |  |  |  |  |
| Secondary | 0.74 (0.55-1.00) | 0.049 | 1.01 (0.93-1.09) | 0.874 |
| Third/higher | 0.80 (0.58-1.10) | 0.177 | 1.04 (0.96-1.13) | 0.360 |
| **Number of conditions** | 1.47 (1.31-1.64) | <0.001 | 1.08 (1.05-1.11) | <0.001 |
| **Complex multimorbidity<sup>b</sup>** | 0.76 (0.52-1.10) | 0.148 | 0.98 (0.90-1.08) | 0.733 |
| **Number of Prescription Medicines (Ref: 2-3 medicines)** |  |  |  |  |
| 0 medicines | 0.01 (0.01-0.01) | <0.001 | 0.94 (0.81-1.08) | 0.371 |
| 1 medicine | 0.36 (0.26-0.49) | <0.001 | 0.65 (0.60-0.71) | <0.001 |
| 4-5 medicines | 1.18 (0.75-1.85) | 0.468 | 1.44 (1.33-1.56) | <0.001 |
| ≥6 medicines | 0.73 (0.46-1.14) | 0.166 | 1.89 (1.73-2.06) | <0.001 |
| **Urban<sup>b</sup>** | 1.02 (0.83-1.24) | 0.876 | 1.09 (1.03-1.16) | 0.002 |
| **Partnered<sup>b</sup>** | 1.06 (0.84-1.33) | 0.639 | 1.03 (0.96-1.10) | 0.382 |
| Intercept | 22.08 (6.35-76.85) | <0.001 | 81.27 (58.48-112.95) | <0.001 |

<sup>a</sup>Model b includes only those with any out-of-pocket prescription medicine expenditure
Note: The variables are mutually adjusted.
<sup>b</sup>Reference groups where not indicated are no LTI scheme eligibility, no private health insurance, male sex, no complex multimorbidity, not urban, and not partnered.
Abbreviations: CI, confidence interval; GMS, General Medical Services; GP, general practitioner; LTI, Long Term Illness

Among those with expenditure, the analysis found a statistically significant positive association between being neither GMS eligible nor having a GP visit-card and level of out-of-pocket prescription medicine expenditure (Rate ratio: 4.74, 95%CI: 4.37-5.15), when compared to those who are GMS eligible (Table 3, model b). A statistically significant positive association was also found between having a GP visit card and level of out-of-pocket prescription medicine expenditure (Rate ratio: 4.70, 95%CI: 4.25-4.37), when compared to those who are GMS eligible. The estimated mean expenditure for those who were GMS eligible was €124 (95%CI: €116-€131). For those with a GP visit-card it was €567 (95%CI: €507-€628) and for those who were neither GMS eligible nor had a GP visit-card it was €547 (95%CI: €508-€586).

No statistically significant association was found between eligibility for the LTI scheme and level of out-of-pocket prescription medicines expenditure (Rate ratio: 0.98, 95%CI: 0.87-1.09). The analysis found a statistically significant positive association between number of chronic conditions and level of out-of-pocket prescription medicine expenditure (Rate ratio: 1.08, 95%CI: 1.05-1.11). The results of the multivariable logit regression and multivariable generalised linear model, with number of chronic conditions included as a categorical variable for sensitivity analysis, were similar to the main analysis (see Appendix A, eTable 2).

### 3.2 Cost-related non-adherence

Overall, 1.7% (n=89) of the sample reported cost-related non-adherence in the previous 12 months. Those experiencing cost-related non-adherence had higher median expenditure on medicines (€240, IQR: €120-€540) than those not reporting cost-related non-adherence (€150, IQR: €30-€360). There were similar levels of cost related non-adherence for those with GMS entitlements (1.7%, n=42) when compared to those with a GP visit card (1.7%, n=10) and those with neither entitlement (1.6%, n=36) (full cost-related non-adherence details in Table 4).

**Table 4.** Out-of-pocket (OOP) medicine expenditure and cost-related non-adherence (CRNA)

|  | **Total % (N)** | **Mean OOP**<br>**Medicine**<br>**Expenditure (SD)** | **Median OOP**<br>**Medicine**<br>**Expenditure (IQR)** |
| --- | --- | --- | --- |
| Proportion reporting any OOP prescription medicine expenditure (N) | 73.4%<br>(4,158) | €434 (504) | €240 (€120-€480) |
| **Cost-related non-adherence** |  |  |  |
| Reporting CRNA in previous 12 months | 1.7% (89) | €393 (€464) | €240 (€120-540) |
| Not reporting CRNA in previous 12 months | 98.3%<br>(5,252) | €336 (€480) | €150 (€30-360) |
| **Entitlements and CRNA** |  |  |  |
| Reporting CRNA with GMS entitlements | 1.7% (42) | €201.14 (€192) | €180 (€120-240) |
| Reporting CRNA with GP visit card entitlements | 1.7% (10) | €662.4 (€634) | €480 (€144-960) |
| Reporting CRNA with neither entitlement | 1.6% (36) | €553.67 (€546) | €450 (€48-780) |
| **No. Chronic Conditions and CRNA** |  |  |  |
| Reporting CRNA with 0 conditions | 0.9% (7) | €27.43 (€73) | €0 (€0-0) |
| Reporting CRNA with 1 condition | 1.3% (17) | €329.65 (€336) | €240 (€60-540) |
| Reporting CRNA with 2 conditions | 1.6% (21) | €367.14 (€539) | €240 (€132-360) |
| Reporting CRNA with 3+ conditions | 2.3% (44) | €488.59 (€479) | €300 (€144-720) |
| **Complex multimorbidity and CRNA** |  |  |  |
| Reporting CRNA with complex multimorbidity | 2.7% (3) | €400 (€400) | €300 (€60-840) |
| Reporting CRNA not with complex multimorbidity | 1.6% (86) | €393 (€468) | €240 (€120-540) |

In the exploratory univariate logit models (Appendix A, eTable 5), statistically significant associations were identified between cost-related non-adherence and number of chronic conditions (OR: 1.21, 95%CI: 1.07-1.36), complex multimorbidity (OR: 1.96, 95%CI: 1.29-2.98) and being on 4-5 medications (compared to 3 medications) (OR: 1.78, 95%CI: 1.02-3.12). However, these results should be interpreted with caution because in the multivariable model, no statistically significant associations were identified (Table 5). Sensitivity analysis, for the multivariable model, with number of chronic conditions included as a categorical variable (Appendix A, eTable 4) yielded similar results.

**Table 5.** Model assessing associations with cost-related non-adherence (CRNA)

|  | Reporting CRNA |  |
| --- | --- | --- |
|  | Odds ratio (95%CI) | p value |
| **Any out-of-pocket prescription medicine expenditure** | 0.82 (0.35-1.91) | 0.644 |
| **Out-of-pocket prescription medicine expenditure (per €100)** | 1.00 (0.94-1.06) | 0.869 |
| **Any out-of-pocket healthcare (excl. prescription medicines) expenditure** | 1.31 (0.71-2.43) | 0.383 |
| **Out-of-pocket healthcare (excl. prescription medicines) expenditure (per €100)** | 1.01 (0.98-1.04) | 0.664 |
| **Healthcare entitlements (Ref: GMS scheme)** |  |  |
| GP visit card | 1.40 (0.59-3.31) | 0.446 |
| Neither GMS scheme nor GP visit card | 1.32 (0.62-2.82) | 0.472 |
| **LTI scheme<sup>a</sup>** | 1.15 (0.54-2.47) | 0.711 |
| **Private Health Insurance<sup>a</sup>** | 0.61 (0.35-1.04) | 0.072 |
| **Age (Years)** | 0.98 (0.95-1.01) | 0.286 |
| **Female sex<sup>a</sup>** | 1.02 (0.65-1.60) | 0.919 |
| **Education (Ref: Primary/none)** |  |  |
| Secondary | 1.41 (0.79-2.54) | 0.247 |
| Third/higher | 1.14 (0.59-2.20) | 0.704 |
| **Number of conditions (per condition)** | 1.06 (0.86-1.31) | 0.557 |
| **Complex multimorbidity<sup>a</sup>** | 1.51 (0.78-2.93) | 0.222 |
| **Number of Prescription Medicines (Ref: 2-3 medicines)** |  |  |
| 0 medications | 0.51 (0.19-1.39) | 0.188 |
| 1 medication | 1.03 (0.52-2.02) | 0.937 |
| 4-5 medications | 1.70 (0.95-3.05) | 0.075 |
| ≥6 medications | 0.92 (0.44-1.94) | 0.824 |
| **Urban<sup>a</sup>** | 1.15 (0.75-1.78) | 0.513 |
| **Partnered<sup>a</sup>** | 0.65 (0.41-1.04) | 0.071 |
| Intercept | 0.04 (0.00-0.58) | 0.018 |

Note: The variables are mutually adjusted.
<sup>a</sup>Reference groups where not indicated are no LTI scheme eligibility, no private health insurance, male sex, no complex multimorbidity, not urban, and not partnered.
Abbreviations: CI, confidence interval; GMS, General Medical Services; GP, general practitioner; LTI, Long Term Illness

## 4. Discussion

### 4.1 Summary

Those with entitlements to heavily publicly-subsidised prescription medicines had much lower out-of-pocket prescription medicine expenditure than those without those entitlements, clearly showing the strong protective financial effects of the GMS scheme in Ireland in 2016. The estimated mean expenditure for those who were GMS eligible was €124, compared to €567 for those with a GP visit-card and €547 for those with neither entitlement. Adjusting for other factors, this corresponded to more than 4.5 times higher out-of-pocket expenditure on prescription medicines for those without GMS eligibility. Given that GMS entitlements are more common among low-income groups, the strong protective financial effects are likely to be particularly beneficial.^43^ No association was seen between LTI scheme entitlement and out-of-pocket prescription medicine expenditure. However, if the LTI scheme did not exist in Ireland, then expenditure in this group would likely be far higher because of the high costs of some medicines covered by the scheme e.g. diabetes medicines.^44^ Therefore, the LTI scheme may be acting as an equaliser for those with conditions covered by the scheme.

The number of chronic conditions an individual has was found to be associated with increased out-of-pocket prescription medicine expenditure. People with zero diagnosed chronic conditions, on average, spent €111 annually on prescription medicines, however this is likely skewed by some with high spending as notably, more than 50% of people with zero diagnosed chronic conditions reported spending nothing out-of-pocket on prescription medicines. For people with two conditions, average annual expenditure was €374, and €432 for those with three or more. The finding of increased prescription medicine expenditure for those with multimorbidity is consistent with studies from a range of contexts.^16,45^

When compared to Denmark, expenditure in for people with multimorbidity is lower than Ireland. An analysis of Danish expenditure in 2020 found that adults with two chronic conditions spent €187 and those with no conditions spent €44.^45^ Data from Korea (adjusted to 2015 values using European Central Bank figures) also showed lower expenditure; average annual expenditure for those aged ≥20 years with three or more chronic conditions was €234.^16^ Conversely, when compared to data from Canada and the US (adjusted to 2015 values using European Central Bank figures),^16^ we find higher out-of-pocket prescription medicine expenditure than for those with multimorbidity in Ireland. In Canada average annual expenditure for those with multimorbidity was €562 among people <65 years and €762 among those ≥65 years.^16^ In the US, average annual expenditure for adults aged <65 years with two chronic conditions was €660, and €882 for those with three conditions.^16^

In relation to cost-related non-adherence, only 1.7% of participants had not filled a prescription in the previous 12 months because of cost. This potentially shows the positive effects of the financial protection measures like the GMS scheme. However, it may also be that non-adherence is underestimated. A 2021/2022 analysis of cost-related medication non-adherence amongst people in Ireland aged ≥40 years with one or more chronic condition found that 16% of participants reported not buying a medication (either prescription or non-prescription) in the previous 12 months because of cost.^46^ This is particularly striking given that payment caps had been significantly lowered by 2021/2022, though the difference might, in large part, be accounted for by the inclusion of non-prescription medicines in the 2021/2022 data. When compared to a 2014 analysis of cost-related non-adherence to prescription medications in those aged ≥55 years in 11 high-income countries,^22^ our estimate from the present study would have ranked second lowest among countries. France was the lowest; 1.6% of participants reported cost-related non-adherence to prescription medications in the previous 12 months.^22^ Whereas 3.1% of UK participants reported cost-related non-adherence despite having much lower prescription charges.^22^ In the multivariable logit regression no statistically significant associations were found between any included variables and cost-related non-adherence. However, in the univariable analysis, number of chronic conditions, complex multimorbidity and being on 4-5 medications (compared to 3 medications) were positively associated with cost-related non-adherence.

### 4.2 Implications and further research

One of the primary mechanisms to reduce out-of-pocket prescription medicine expenditure is provision of healthcare entitlements. Given the significant protective effects of the GMS scheme, ensuring full uptake amongst those eligible would be beneficial, as a 2021 study found that 31% of those with an entitlement to the GMS scheme did not have a GMS card.^47^ In relation to improving entitlements, since 2016, DPS and GMS payment caps have been progressively lowered.^48,49^ Consideration is being given to further changes,^50,51^ such as introducing a lower DPS cap for single-headed households or lowering the fee per item for those with GMS entitlements.^13^ The Irish government have explicitly stated that they do not plan to extend the LTI scheme to other conditions.^52^ In relation to welfare entitlement thresholds, organisations such as the Irish Human Rights and Equality Commission have suggested that these thresholds should increase in line with inflation.^53^ However, there has been limited expansion of entitlements to the GMS scheme,^54^ which provides maximal protection. Instead, GP visit cards entitlements have been extended,^55^ however this group have no additional protection from out-of-pocket prescription medicine expenditure. There may be barriers to increasing healthcare entitlements as, even without any changes to entitlements, the healthcare budget is already expected to take up a greater proportion of the overall state budget due to various factors including the ageing population.^56^ The healthcare budget may also face constraints due to higher public spending on pharmaceuticals than the EU average.^37^

There are other initiatives that can reduce out-of-pocket prescription medicine expenditure. Prescription medicine prices are higher in Ireland than many other countries.^57^ Various mechanisms could reduce costs for the patient, including increased generic medicine use promoted by reference pricing and encouraging or mandating prescribing by International Nonproprietary Name.^58,59^ Also, fixed mark-ups^60^ or mandated price transparency (to increase competition)^61^ on prescription medicines in community pharmacies could reduce costs. However, supply side interventions may have a larger impact on prescription medicine prices.^62^ Examples of this might include shorter monopolies on patented drugs, or limiting the ability of manufacturers to extend patents.^62^ Deprescribing interventions should also be considered, where a healthcare professional evaluates a patient’s regular medicines for any that are unnecessary or inappropriate and can therefore be reduced or stopped.^63^ This can positively affect health outcomes, and reduce out-of-pocket medicine expenditure.^63^ Cost-of-care conversations (a healthcare professional and patient discussing out-of-pocket healthcare costs and entitlements)^64^ could also reduce expenditure for patients^65^ and improve clinical outcomes (though greater adherence).^66^

Given the Sláintecare aim of universal access to care and current proposed changes to several prescription medicines entitlements,^13,50,53,67^ future research modelling the effects of different entitlement changes would provide valuable evidence. Analysing out-of-pocket prescription medicine expenditure for those <50 years would be valuable as younger populations are more likely to experience cost related non-adherence.^31^ The findings of this study that cost-related non-adherence is low may imply that people are making sacrifices in other areas of their life to afford their healthcare,^68,69^ which could be evaluated in future research.

### 4.3 Strengths and Limitations

A strength of this study is the nationally representative sample captured by TILDA. In terms of limitations, self-report is associated with recall bias. For example, it can lead to under-reporting of chronic conditions,^70,71^ which may partly be the reason for the somewhat high out-of-pocket prescription medicine expenditure found for those with zero chronic conditions. Though this may also attributable to medicines for acute conditions or for chronic conditions not captured by TILDA (e.g. menopause and dermatological conditions). There is mixed evidence of the accuracy of self-report for medicine use.^72,73^ Though notably, self-report data from TILDA was found to be accurate for services such as the general practitioner and the outpatient department.^74^ Scaling of reported monthly expenditure to estimate annual out-of-pocket prescription medicine expenditure may not have accurately captured annual expenditure, though the shorter recall period likely reduces recall bias.^75^ Another limitation is that cost-related non-adherence may have been underestimated, possibly because of the highly specific phrasing of the question referring to prescriptions from one’s GP. A more comprehensive and dynamic measure of non-adherence or unmet need may be necessary to accurately capture the issues at play.^76^ The significant exploratory univariate findings for cost-related non-adherence may indicate that the multivariable logit regression is underpowered, or that these associations are partly explained by other factors adjusted for. Measuring financial burden as the percentage of equivalised household income spent on prescription medicines could be considered a limitation. A more valid measure would have been the proportion of non-subsistence income (income remaining after expenditure on subsistence) one spends on healthcare.^77^ However, this is not available from TILDA.

### 4.4 Conclusion

The entitlements system in Ireland offers significant protection against out-of-pocket prescription medicine expenditure for approximately a third of the population with GMS entitlements, which is mostly those on low incomes. Among those who do not have these protections, out-of-pocket prescription medicine expenditure is much higher, with people who have multimorbidity most at risk of high out-of-pocket prescription medicine expenditure. However, cost-related non-adherence appears to be low. A range of measures could be introduced to reduce out-of-pocket prescription medicine expenditure including increased entitlement thresholds, increased generic prescribing and initiatives to increase the uptake of entitlements, especially for those with multimorbidity

## Data Availability

Researchers interested in using TILDA data can find out details about how to access it here: https://tilda.tcd.ie/data/accessing-data/

