## Appendix A for "Out-of-pocket prescription medicine expenditure amongst community-dwelling adults: findings from The Irish Longitudinal Study on Ageing (TILDA) in 2016"

### eBox 1. Healthcare Coverage in Ireland

At the time of data collection for the present study (2016), those with public healthcare entitlements under the General Medical Services (GMS) scheme paid €2.50 per prescribed item, and this expenditure was capped at €25 per household per month.^22^ The GMS scheme is means tested based on income and household structure, with different income thresholds for different age groups. For example, the GMS scheme income threshold for a single person aged 65 years or under is €184 a week (after taxes and allowable expenses).^23^ GMS eligibility can also be provided on a discretionary basis e.g. for severe illness.^24^ For those who are not eligible for the GMS scheme, individuals pay the full price of medications out-of-pocket, but payments are capped at €144 per household per month (during the study period) with the Drugs Payment Scheme (DPS) covering costs in excess of this. ^25^

Another major entitlement provision in Ireland is the Long-Term Illness (LTI) scheme which entitles individuals with any one of sixteen specified health conditions (e.g. diabetes, cerebral palsy) to free prescribed medicines to treat that condition.^26^ The LTI scheme is not means tested.^27^ There is less clarity around the medication subsidies provided by private health insurance^28,29^ though they are likely to be limited as most insurance plans are hospital plans.^30^ Other relevant schemes include the *GP Visit Card* which entitles holders to free access to general practitioners (GPs) but provides no specific cover for medicines,^31^ and the Health Amendment Act which entitles those affected by contaminated blood products provided by the State to a similar range of healthcare services to those eligible for the GMS scheme.^32^ The High Tech Drug Scheme entitles those on the LTI scheme, GMS scheme or with Health Amendment Act eligibility to access *high tech* medicines free of charge.^33^ For people on the drugs payment scheme they pay for high tech drugs up to the €144 monthly threshold. The high tech drugs scheme only applies to *high tech* medicines prescribed by a hospital consultant.^33^ Finally, there is also significant tax relief available for health expenses, such as out-of-pocket payments for medicines.^34^

### eBox 2. Medicine questions in TILDA wave 4

**HU035: Not counting health insurance refunds, on average about how much [do/does] [you/he/she] pay out-of-pocket for [your/his/her] prescribed drugs per month?**

IWER: IF RESPONDENT DOES NOT PURCHASE PRESCRIBED DRUGS REGULARLY, ASK FOR TOTAL SPENT IN THE LAST 12 MONTHS IN PRESCRIBED DRUGS AND DIVIDE BY 12.

IWER: IF R CANNOT GIVE EXACT VALUE, ACCEPT APPROXIMATE VALUE

VALUE [Allow Responses up to 2 decimal places]

€0.00 … €5,000.00

-98. DK

-99. RF

NOTE: Include the €2.50 per prescribed item charges for medical card holders.

NOTE: Do not consider expenses for self-medication or drugs not prescribed

NOTE: By ‘out of pocket’ expenses we mean everything that is not paid by the insurance company. If [you/he/she] first [pay/pays] but later get it refunded, this is not out of pocket expenses.

(SHARE)

**MD001a:** **Now I would like to record all medications that [you/Rname] [take/takes] on a regular basis, like every day or every week. This will include prescription and non-prescription medications, over-the-counter medicines, vitamins, and herbal and alternative medicines.**

**Do/Does [you/Rname] take any medication on a regular basis?**

1. Yes	**GO TO MD001**

5. No

98. DK

99. RF

**IF INTSTATUSW4=1,2 OR 3 & MD001A = 5 GO TO MD008**

**IF INTSTATUSW4= 4 OR 5 & MD001A = 5 GO TO NEXT SECTION**

**IWER: ASK RESPONDENT IF YOU COULD SEE THE MEDICATIONS HE/SHE TAKES SO YOU CAN COPY DOWN THE CORRECT SPELLING OF EACH TABLET.**

**IWER: PROMPT: DO I HAVE ALL OF [YOUR/RName’s] MEDICATIONS HERE?**

**DISPLAY NOTE: INPUT THE BRAND NAME WHEN AVAILABLE, RATHER THAN THE GENERIC NAME FOR ANY MEDICATIONS**

**MD001: TYPE THE FIRST FOUR LETTERS OF THE MEDICATION. YOU WILL BE GIVEN A LIST OF POSSIBILITIES CHOOSE ONE. Each medication is recorded in a separate variable**

**(MD001_01 – MD001_20; MD001_ATC_01 - MD001_ATC_20; MD001_NONPROP_01 - MD001_NONPROP_20)**

95. Other (specify) GO TO MD001oth

98. DK

99. RF

**MD001oth: Other (specify)**

**IF THE MEDICATION DOES NOT APPEAR ON THE LIST CAREFULLY TYPE THE FULL MEDICATION NAME. MAKE SURE YOU TYPE THE NAME OF THE BRAND USED AND NOT THE CHEMICAL NAME.**

**(MD001OTH_01-MD001OTH_20)**

Text: up to 60 characters GO TO MD005

**MD005: Was this medication prescribed by a doctor or did you get it over the counter? (MD005_01-MD005_20)**

1. Prescribed by a doctor

2. Over the counter

98. DK

99. RF

**(Note to Scripters -** For medications that are available over the counter as well as on prescription, we are interested in how the respondent got them.)

**RETURN AND REPEAT MD001 FOR UP TO 20 MEDICATIONS PER PERSON**

**IF instatusW4= 4 OR 5 GO TO NEXT SECTION**

**MD006: In the last 12 months have you ever forgotten to take any of the medicines you are supposed to regularly take?**

1. Yes GO TO MD007

5. No GO TO MD008

98. DK GO TO MD008

99. RF GO TO MD008

**MD007: How often have you forgotten to take your medication in the last 12 months?**

1. Rarely

2. Sometimes

3. Often

98. DK

99. RF

**MD008: In the last 12 months, have you ever received a prescription from your GP that you didn’t fill with the pharmacy because you thought that the medication was too expensive?**

1. Yes

5. No

98. DK

99. RF

### eBox 3. Prescription medicine expenditure outlier management

Med exp methods

We replaced data with missing for anything above €288 (twice the drugs payment scheme (DPS) limit), unless there is other information provided by the participant to suggest it is plausible or a specific error has been made, e.g.

•	If they have a medical card, report a multiple of €250, and their number of prescription medicines is equivalent (+/- 1) to the number of medicines that would give that charge (e.g. €1250 and reporting 4-6 meds)

•	If not on the General Medical Services (GMS) scheme (so likely using DPS) and report a multiple of 12 (i.e. giving a whole number if dividing by 12), then assume to be an annual rather than monthly report.

•	If they remain as an outlier, checked if any of the medicines they report are not covered by the DPS.

### eBox 4. Health coverage questions

**HU001: [Is/Are] [you/he/she] covered by:**

IWER: CODE THE ONE THAT APPLIES

1. Full Medical Card or equivalent

2. GP Visit Card

96. Neither of these

98. DK

99. RF

**Note:** This question is asked even of those covered by private medical insurance. Most over 70s are entitled to medical cards.

(EU-SILC)

**HU070: [Is/Are] [you/he/she] covered by:**

1. The long term illness scheme
2. A Health Act Amendment Card

96. Neither of these

98 DK

99 RF

**HU002: [Do/Does] [you/he/she] have private medical insurance cover (VHI etc.) in [your/his/her] own name or through another family member?**

1. Yes, in own name GO TO HU003

2. Yes, as the spouse of a subscriber GO TO HU003

3. Yes, as the relative of a subscriber GO TO HU003

5. No	GO TO HU049

98. DK	GO TO HU049

99. RF	GO TO HU049

(HEALTH INSURANCE AUTHORITY 2005 SURVEY)

### eBox 5. Question about health conditions in TILDA

(A) Since your last interview, has a doctor ever told you/ that you have any of the [other] conditions on this card?

(B) Has a doctor ever told [you/Rname] that [you/he/she] [have/has] any of the conditions on this card?

**IWER: PROBE - 'WHAT OTHERS?' CODE ALL THAT APPLY.**

1. Chronic lung disease such as chronic bronchitis
or emphysema	**GO TO PH302	[ph301_01]**

2. Asthma	**[ph301_02]**

3. Arthritis (including osteoarthritis, or rheumatism)	**GO TO PH304	[ph301_03]**

4. Osteoporosis, sometimes called thin or brittle bones	**[ph301_04]**

5. Cancer or a malignant tumour	**GO TO PH309	[ph301_05]**

(including leukaemia or lymphoma but excluding minor skin cancers)

6. Parkinson's disease	**GO TO PH314	[ph301_06]**

7. Any emotional, nervous or psychiatric problems,

such as depression or anxiety	**GO TO PH315	[ph301_07]**

9. Alzheimer's disease	**GO TO PH318	[ph301_09]**

10. Dementia, organic brain syndrome, senility	**GO** **TO PH319**	**[ph301_10]**

11. Serious memory impairment	**GO TO PH319a	[ph301_11]**

12. Stomach ulcers	**[ph301_12]**

13. Varicose Ulcers (an ulcer due to varicose veins)	**[ph301_13]**

14. Cirrhosis, or serious liver damage	**[ph301_14]**

15. Thyroid Problems	**GO TO PH325	[ph301_15]**

16. Alcohol abuse	**GO TO PH320a	[ph301_16]**

17. Substance abuse	**GO TO PH320b	[ph301_17]**

18. Chronic kidney disease	**GO TO PH327	[ph301_18]**

19. Severe Anaemia	**[ph301_19]**

20. Epilepsy	GO TO PH329	**[ph301_20]**

95. Other (please specify)	**[ph301_95] [ph301oth]**

96. None of these	**GO TO PH328	[ph301_96]**

98. DK	**GO TO PH328	[ph301_98]**

99. RF	**GO TO PH328	[ph301_99]**

(ELSA/ similar question HRS/NSHAP)

**IWER: SHOW CARD PH2 [PAGE #]**

**IF (intstatusW4 = 2, 4, 5), USE WORDING ‘B’. ALL OTHERS, USE WORDING ‘A’.**

PH201: Please look at card PH2.

(A) Since your last interview, has a doctor ever told you that you have any of the [other] conditions on this card?

(B) Has a doctor ever told [you/Rname] that [you/he/she] [have/has] any of the conditions on this card?

INTERVIEWER: PROBE - 'WHAT OTHERS?' CODE ALL THAT APPLY.

1. High blood pressure or hypertension	**[ph201_01]**

2. Angina	**[ph201_02]**

3. A heart attack

(including myocardial infarction or coronary thrombosis)	**[ph201_03]**

4. Congestive heart failure	**[ph201_04]**

5. Diabetes or high blood sugar	**[ph201_05]**

6. A stroke (cerebral vascular disease)	**[ph201_06]**

7. Ministroke or TIA	**[ph201_07]**

8. High cholesterol	**[ph201_08]**

9. A heart murmur	**[ph201_09]**

11. Atrial Fibrillation	**[ph201_11]**

12. An abnormal heart rhythm (not atrial fibrillation)	**[ph201_12]**

95. Any other heart trouble (please specify)	**[ph201_95][ph201oth]**

96. None of these	**[ph201_96]**

98. DK	**[ph201_98]**

99. RF	**[ph201_99]**

(ELSA/ similar questions in HRS/ SHARE)

(A) Since [your] last interview, has a doctor ever told you that you have any of the following [other] eye diseases?

(B) Has a doctor ever told [you/Rname] that [you/he/she] [have/has] any of the following eye diseases? [DISPLAY ALL CONDITIONS]

**IWER: READ OUT. CODE ALL THAT APPLY.**

1. Cataracts	**[ph105_01]**

2. Glaucoma	**[ph105_02]**

3. Age related macular degeneration	**[ph105_03]**

95. Other (please specify)	**[ph105_95] [ph105oth]**

### eBox 6. Conditions included in analysis (partly based on Ryan and colleagues)

1. Cardiac Condition

- Angina
- Heart Attack
- Congestive Heart Failure
- Heart Murmur
- Abnormal heart rhythm
- Atrial fibrillation
- Any heart trouble

2. Cerebrovascular disease

- Stroke
- TIA

3. HTN
4. Diabetes
5. High Cholesterol
6. Chronic respiratory disease

- Chronic lung disease
- Asthma

7. Liver disease

- Alcohol abuse
- Cirrhosis
- Liver disease

8. Eye disease

- Cataracts
- Glaucoma
- Age related macular degeneration
- Other eye disease

9. Cognitive Impairment

- Alzheimer’s
  - Dementia
  - Serious cognitive impairment

10. Arthritis
11. Osteoporosis
12. Cancer
13. Parkinson’s disease
14. Emotional /psychological condition including anxiety and depression
15. Substance abuse
16. Stomach Ulcers
17. Varicose veins including varicose ulcers
18. Epilepsy
19. Thyroid Problems
20. Chronic kidney disease
21. Anaemia

### eBox 7. International Classification of Primary Care (ICPC) Classification of Body Systems

1. Cardiovascular
2. Respiratory
3. Digestive
4. Eyesystem
5. Psychological
6. Metabolic
7. Musculoskeletal
8. Neurological
9. Urological

### eBox 8. Healthcare Expenditure Questions

**HU005: In the last 12 months, about how often did [you/he/she] visit [your/his/her] GP?**

IWER: IF RESPONDENT HAS NOT VISITED GP IN THE LAST 12 MONTHS CODE **0**

0…200

-98. DK	GO TO HU007

-99. RF	GO TO HU007

IF (HU005=0, -98,-99) GO TO HU007

IF (HU005>0 AND HU001=1, 2) GO TO HU007

IF (HU005>0 AND HU070= 2) GO TO HU007

IF (HU005>0 AND HU001≠1, 2) GO TO HU006

IF (HU005>0 AND HU070 ≠ 2) GO TO HU006

(SHARE)

**HU006: How much did [you/he/she] pay for [your/his/her] last visit to the GP, after any health insurance reimbursement?’**

IWER: IF R CANNOT GIVE EXACT VALUE, ACCEPT APPROXIMATE VALUE

€0.00 … €10,000

-98. DK

-99. RF

**HU038: In total, how much did [you/he/she] pay for all of [your/his/her] A&E visit(s) in the last 12 months, after any health insurance reimbursement?’**

IWER: IF R CANNOT GIVE EXACT VALUE, ACCEPT APPROXIMATE VALUE

€0 … €10,000

-98. DK

-99. RF

**HU075: In total, how much did [you/he/she] pay for [your/his/her] outpatient/day patient visits in the last 12 months, after any health insurance reimbursement?** (May be zero)

IWER: IF R CANNOT GIVE EXACT VALUE, ACCEPT APPROXIMATE VALUE

€0 … €50,000

-98. DK

-99. RF

If HU062=1

**HU039: In total, how much did [you/he/she] pay for [your/his/her] visit(s) to consultant(s) in the last 12 months, after any health insurance reimbursement?** (May be zero)

IWER: IF R CANNOT GIVE EXACT VALUE, ACCEPT APPROXIMATE VALUE

€0 … €20,000

-98. DK

-99. RF

**HU040: In total, how much did [you/he/she] pay for [your/his/her] overnight hospital stays(s) in the last 12 months, after any health insurance reimbursement?** (May be zero)

IWER: IF R CANNOT GIVE EXACT VALUE, ACCEPT APPROXIMATE VALUE

€0 … €50,000

-98. DK

-99. RF

**HOME CARE**

**IF HH002=1 GO TO HU076**

**IF HH002=2, 3 GO TO HU083**

**HU015_a4: Not counting costs paid by the HSE/health board, about how much did [you/Rname] (and [your/his/her] [husband/wife/partner]) pay for this home help in the last month?** (May be zero)

€0 … €10,000

-98. DK

-99. RF

GO TO HU015

**HU015_b4: Not counting costs paid by the HSE/health board, about how much did [you/Rname] (and [your/his/her] [husband/wife/partner]) pay this personal care attendant in the last month?** (May be zero)

€0 … €10,000

-98. DK

-99. RF

GO TO HU015

**HU015_d4: Not counting costs paid by the HSE/health board, about how much did [you/Rname] (and [your/his/her] [husband/wife/partner]) pay for this Home Care Package in the last month?** (May be zero)

€0 … €10,000

-98. DK

-99. RF

GO TO HU015

**HU036: Not counting any refunds from [your/his/her] health insurance, about how much did [you /he/she] pay (out-of-pocket) for any other health expenses [you/he/she] had in the last 12 months?**

€0 … and €20,000

-98. DK

-99. RF

**Note:** By other health expenses we mean non-prescription drugs, private physiotherapy, preventive rehabilitative services such as occupational therapy etc.

By ‘out of pocket’ expenses we mean everything that is not paid by the insurance company. If [you/he/she] first pay/pays] but later [get/gets] it refunded, this is not out of pocket expenses. Prescription drugs should be included in HU035 and not here.

(SHARE)

### eTable 1. Out-of-pocket prescription medicine expenditure for individual conditions

| **Condition** | **% (N)** | **Mean OOP Prescription Medicine Expenditure (SD)** | **Median OOP Prescription Medicine Expenditure (IQR)** |
| --- | --- | --- | --- |
| Hypertension | 37.6% (2,131) | €399 (492) | €240 (120-420) |
| High Cholesterol | 36.8% (2,087) | €390 (502) | €204 (60-456) |
| Arthritis | 35.7% (2,026) | €371 (503) | €180 (60-360) |
| Eye Disease | 17.1% (970) | €378 (513) | €192 (90-324) |
| Osteoporosis | 15.9% (903) | €404 (517) | €240 (90-444) |
| Cardiac Condition | 14.2% (805) | €462 (557) | €240 (120-480) |
| Chronic Respiratory Disease | 11.2% (637) | €526 (594) | €300 (120-720) |
| Thyroid Problems | 9.2% (524) | €431 (482) | €282 (120-540) |
| Diabetes | 9.0% (512) | €292 (448) | €150 (24-300) |
| Emotional/Psychological Condition including Anxiety and Depression | 7.5% (423) | €399 (478) | €240 (120-360) |
| Gastrointestinal Conditions including Stomach Ulcers | 2.9% (163) | €382 (473) | €240 (120-336) |
| Cancer | 2.7% (152) | €447 (568) | €198 (90-540) |
| Varicose Veins including Varicose Ulcers | 1.7% (96) | €302 (399) | €180 (87-300) |
| Cerebrovascular Disease | 1.2% (69) | €516 (717) | €204 (90-480) |
| Chronic Kidney Disease | 0.8% (44) | €452 (519) | €144 (0-312) |
| Liver Disease | 0.7% (42) | €354 (459) | €240 (72-360) |
| Parkinson’s Disease | 0.6% (36) | €417 (640) | €222 (15-360) |
| Cognitive Impairment | N.A. (<30) | N.A. | N.A. |
| Epilepsy | N.A. (<30) | N.A. | N.A. |
| Substance Abuse | N.A. (<30) | N.A. | N.A. |
| Anaemia | N.A. (<30) | N.A. | N.A. |

### eTable 2. Logit and generalised linear models assessing associations with out-of-pocket (OOP) prescription medicine expenditure (chronic conditions included as categorical variable)

|  | Any OOP expenditure |  | Value of OOP expenditure^a^ |  |
| --- | --- | --- | --- | --- |
|  | Odds ratio (95%CI) | p value | Rate ratio (95%CI) | p value |
| **Healthcare entitlements (Ref: GMS scheme)** |  |  |  |  |
| GP visit card-holder | 0.80 (0.55-1.19) | 0.273 | 4.67 (4.23-5.16) | <0.001 |
| Neither GMS scheme nor GP visit card | 0.62 (0.47-0.83 | 0.001 | 4.72 (4.35-5.13) | <0.001 |
| **LTI scheme**^b^ | 0.67 (0.46-0.98) | 0.037 | 0.98 (0.88-1.10) | 0.788 |
| **Private Health Insurance**^b^ | 1.09 (0.85-1.41) | 0.506 | 1.07 (0.99-1.15) | 0.074 |
| **Age (years)** | 1.00 (0.99-1.02) | 0.793 | 1.01 (0.93-1.09) | 0.008 |
| **Female sex**^b^ | 0.77 (0.62-0.94) | 0.012 | 0.92 (0.86-0.98 | 0.006 |
| **Education (Ref: primary/none)** |  |  |  |  |
| Secondary | 0.75 (0.55-1.01) | 0.055 | 1.01 (0.93-1.09) | 0.832 |
| Third/higher | 0.81 (0.59-1.11) | 0.184 | 1.04 (0.95-1.13) | 0.388 |
| **Number of conditions (ref: 0 conditions)** |  |  |  |  |
| 1 | 2.09 (1.59-2.76) | <0.001 | 1.04 (0.92-1.17) | 0.571 |
| 2 | 3.01 (2.21-4.11) | <0.001 | 1.13 (1.00-1.28) | 0.045 |
| 3+ | 3.58 (2.28-5.63) | <0.001 | 1.27 (1.10-1.46) | 0.001 |
| **Complex multimorbidity**^b^ | 1.12 (0.73-1.72) | 0.610 | 1.02 (0.92-1.13) | 0.678 |
| **Number of Prescription Medicines (Ref: 2-3 medicines)** |  |  |  |  |
| 0 medicines | 0.01 (0.01-0.01) | <0.001 | 0.94 (0.81-1.09) | 0.403 |
| 1 medicine | 0.36 (0.27-0.50) | <0.001 | 0.65 (0.60-0.71) | <0.001 |
| 4-5 medicines | 1.22 (0.78-1.91) | 0.381 | 1.44 (1.33-1.56) | <0.001 |
| ≥6 medicines | 0.81 (0.51-1.27) | 0.358 | 1.93 (1.76-2.10) | <0.001 |
| **Urban**^b^ | 1.02 (0.83-1.25) | 0.852 | 1.10 (1.04-1.16) | 0.001 |
| **Partnered**^b^ | 1.05 (0.83-1.32) | 0.697 | 1.03 (0.96-1.10) | 0.417 |
| Intercept | 15.79 (4.47-55.72) | <0.001 | 82.02 (58.09-115.80) | <0.001 |

^a^Model b includes only those with any out-of-pocket prescription medicine expenditure

Note: The variables are mutually adjusted

^b^Reference groups where not indicated are no LTI scheme eligibility, no private health insurance, male sex, no complex multimorbidity, not urban, and not partnered.

Abbreviations: CI, confidence interval; GMS, General Medical Services; GP, general practitioner; LTI, Long Term Illness

### eTable 3. Demographic and entitlement characteristics of sample broken down by alternative measures of out-of-pocket prescription medicine expenditure

|  | Proportion of out-of-pocket healthcare expenditure spent on prescription medicines Mean (SD) | Proportion of out-of-pocket healthcare expenditure spent on prescription medicines Median (IQR) | Financial burden  Mean (SD) | Financial burden  Median (IQR) |
| --- | --- | --- | --- | --- |
| Age (years) |  |  |  |  |
| <60 | 51.8% (41.4%) | 61.5% (0.0-95.2) | 2.5% (7.6%) | 0.4% (0.0-2.5) |
| 60-69 | 62.4% (38.6%) | 75.1% (28.6—100.0) | 3.2% (8.0%) | 1.1% (0.0-3.3) |
| 70-79 | 79.2% (32.0%) | 100.0% (64.7-100.0) | 3.9% (10.2%) | 1.6% (0.5-3.8) |
| 80-89 | 85.5% (26.9%) | 100.0% (84.2-100.0) | 5.2% (11.3%) | 2.5% (1.0-5.3) |
| 90+ | 91.9% (20.7%) | 100.0% (100.0-100.0) | 6.6% (15.2%) | 2.2% (0.8-4.4) |
| Sex |  |  |  |  |
| Female | 68.4% (37.7%) | 85.2% (41.9-100.0) | 3.8% (9.4%) | 1.4% (0.0-3.8) |
| Male | 68.4% (38.0%) | 87.0% (40.0-100.0) | 3.3% (8.9%) | 1.1% (0.0-3.1) |
| Education |  |  |  |  |
| Primary/none | 84.9% (29.7%) | 100.0% (87.8-100.0) | 4.2% (10.4%) | 1.7% (0.5-3.8) |
| Secondary | 67.3% (38.4%) | 84.4% (37.5-100.0) | 3.5% (9.0%) | 1.2% (0.0-3.4) |
| Third/higher | 56.7% (38.4%) | 70.6% (21.6-99.6) | 3.2% (8.4%) | 1.0% (0.0-3.3) |
| Area of residence |  |  |  |  |
| Urban | 67.7% (37.3%) | 82.8% (40.0-100.0) | 3.9% (10.2%) | 1.4% (0.0-3.8) |
| Not Urban | 69.1% (38.4%) | 90.9% (41.2-100.0) | 3.2 (7.9%) | 1.2% (0.0-3.2) |
| Marital Status |  |  |  |  |
| Partnered | 64.7% (38.5%) | 78.9% (33.3-100.0) | 3.0% (7.8%) | 1.1% (0.0-3.1) |
| Not Partnered | 76.5% (34.9%) | 100.0 (58.3-100.0) | 4.7% (11.5%) | 1.8% (0.3-4.5) |
| Private Health Insurance |  |  |  |  |
| Yes | 57.5% (37.8%) | 66.7% (23.1-95.6) | 3.7% (9.5%) | 1.2% (0.0-3.7) |
| No | 86.1% (30.6%) | 100.0% (99.7-100.0) | 3.3% (8.6%) | 1.4% (0.2-3.3) |
| LTI Scheme |  |  |  |  |
| Yes | 71.9% (37.9%) | 100.0% (46.2-100.0) | 4.2% (11.5%) | 1.3% (0.1-3.7) |
| No | 68.1% (37.8%) | 85.4% (40.0-100.0) | 3.5% (8.9%) | 1.3% (0.0-3.5) |
| GMS & GP Entitlements |  |  |  |  |
| GMS Scheme | 87.4% (27.4%) | 100.0% (100.0-100.0) | 3.1% (9.1%) | 1.4% (0.4-3.0) |
| GP Visit Card-Holder | 79.0% (28.7%) | 91.5% (66.7-100.0) | 6.0% (9.7%) | 3.1% (1.0-7.4) |
| Neither | 45.4% (37.0%) | 54.4% (0.0-79.2) | 3.4% (9.0%) | 0.8% (0.0-3.4) |
| Number of Prescription Medicines (quintiles) |  |  |  |  |
| 0 regular medicines | 16.7% (34.8%) | 0.00 (0.00-0.00) | 0.4% (1.6%) | 0.0% (0.0-0.0) |
| 1 regular medicine | 68.9% (33.6%) | 78.3% (44.4-100.0) | 2.2% (6.1%) | 0.8% (0.3-2.1) |
| 2-3 regular medicines | 78.6% (27.7%) | 93.4% (63.2-100.0) | 4.2% (10.4%) | 1.7% (0.8-3.9) |
| 4-5 regular medicines | 83.9% (24.2%) | 100.0 (72.7-100.0) | 5.3% (9.9%) | 2.8% (1.4-5.5) |
| 6+ regular medicines | 87.4% (22.5%) | 100.0% (82.5-100.0) | 7.4% (13.4%) | 4.0% (2.0-7.2) |
| Number of Chronic Conditions |  |  |  |  |
| 0 chronic conditions | 37.5% (44.0%) | 0.0% (0.0-92.3) | 1.2% (5.7%) | 0.0% (0.0-0.5) |
| 1 chronic condition | 62.5% (39.1%) | 75.0 (28.6-100.0) | 2.4% (6.2%) | 0.7% (0.0-2.5) |
| 2 chronic conditions | 72.8% (34.2%) | 88.9% (53.7-100.0) | 4.0% (10.1%) | 1.5% (0.5-3.7) |
| 3+ chronic conditions | 80.2% (29.3%) | 100.0 (65.8-100.0) | 5.2% (11.1%) | 2.5% (1.0-5.3) |
| Complex Multimorbidity |  |  |  |  |
| Yes | 80.6% (28.8%) | 100.0% (23.5-100.0) | 5.4% (11.4%) | 2.6% (1.1-5.5) |
| No | 62.8% (40.1%) | 78.3% (23.5-100.0 | 2.8% (7.9%) | 0.8% (0.0-2.6%) |

### eTable 4. Model assessing associations with cost-related non-adherence (CRNA) with chronic conditions included as categorical variable

|  | Reporting CRNA |  |
| --- | --- | --- |
|  | Odds ratio (95%CI) | p value |
| **Any out-of-pocket prescription medicine expenditure (Ref: no out-of-pocket prescription medicine expenditure)** | 0.82 (0.35-1.92) | 0.653 |
| **Out-of-pocket prescription medicine expenditure** | 1.00 (1.00-1.00) | 0.524 |
| **Non-prescription medicine out-of-pocket expenditure** | 1.00 (1.00-1.00) | 0.515 |
| **Healthcare entitlements (Ref: GMS scheme)** |  |  |
| GP visit card-holder | 1.45 (0.62-3.41) | 0.391 |
| Neither GMS scheme nor GP visit card-holder | 1.53 (0.78-3.01) | 0.214 |
| **LTI scheme**^a^ | 1.17 (0.55-2.51) | 0.679 |
| **Private Health Insurance**^a^ | 0.64 (0.38-1.09) | 0.101 |
| **Age (years)** | 0.98 (0.95-1.01) | 0.294 |
| **Female sex**^a^ | 1.03 (0.66-1.61) | 0.891 |
| **Education (Ref: primary/none)** |  |  |
| Secondary | 1.43 (0.80-2.57) | 0.229 |
| Third/higher | 1.15 (0.60-2.24) | 0.670 |
| **Number of conditions (Ref: 0 conditions)** |  |  |
| 1 | 1.28 (0.51-3.23) | 0.602 |
| 2 | 1.47 (0.58-3.75) | 0.416 |
| 3+ | 1.26 (0.41-3.91) | 0.684 |
| **Complex multimorbidity**^a^ | 1.82 (0.83-3.99) | 0.137 |
| **Number of Prescription Medicines (Ref: 2-3 medicines)** |  |  |
| 0 medicines | 0.54 (0.19-1.49) | 0.232 |
| 1 medicine | 1.04 (0.53-2.05) | 0.910 |
| 4-5 medicines | 1.72 (0.96-3.08) | 0.069 |
| ≥6 medicines | 0.96 (0.46-2.01) | 0.911 |
| **Urban**^a^ | 1.16 (0.75-1.78) | 0.505 |
| **Partnered**^a^ | 0.65 (0.41-1.04) | 0.073 |
| Intercept | 0.04 (0.00-0.51) | 0.014 |

Note: The variables are mutually adjusted.

^a^Reference groups where not indicated are no LTI scheme eligibility, no private health insurance, male sex, no complex multimorbidity, not urban, and not partnered.

Abbreviations: CI, confidence interval; GMS, General Medical Services; GP, general practitioner; LTI, Long Term Illness

### eTable 5. Univariate logistic model assessing associations with cost-related non-adherence (CRNA)

|  | Reporting CRNA |  |
| --- | --- | --- |
|  | Odds ratio (95%CI) | p value |
| **Any out-of-pocket prescription medicine expenditure** | 1.45 (0.83-2.54) | 0.192 |
| **Out-of-pocket prescription medicine expenditure (per €100)** | 1.02 (0.98-1.06) | 0.265 |
| **Any out-of-pocket healthcare (excl. prescription medicines) expenditure** | 1.13 0.74-1.72) | 0.571 |
| **Out-of-pocket healthcare (excl. prescription medicines) expenditure (per €100)** | 1.01 (0.99-104) | 0.340 |
| **Healthcare entitlements (Ref: GMS scheme)** |  |  |
| GP visit card holder | 1.02 (0.51-2.05) | 0.947 |
| Neither GMS scheme nor GP visit card | 0.99 (0.63-1.55) | 0.961 |
| **LTI scheme**^a^ | 1.23 (0.59-2.57) | 0.573 |
| **Private Health Insurance**^a^ | 0.69 (0.46-1.05) | 0.087 |
| **Age (years)** | 0.99 (0.97-1.02) | 0.574 |
| **Female sex**^a^ | 1.21 (0.79-1.86) | 0.386 |
| **Education (Ref: primary/none)** |  |  |
| Secondary | 1.33 (0.77-2.31) | 0.302 |
| Third/higher | 1.00 (0.56-1.79) | 0.996 |
| **Number of conditions (per condition)** | 1.21 (1.07-1.36) | 0.002 |
| **Number of conditions (ref:0 conditions)** |  |  |
| 1 condition | 1.49 (0.61-3.60) | 0.379 |
| 2 conditions | 1.87 (0.79-4.41) | 0.155 |
| 3+ conditions | 2.61 (1.17-5.82) | 0.019 |
| **Complex multimorbidity**^a^ | 1.96 (1.29-2.98) | 0.002 |
| **Number of Prescription Medicines (Ref: 2-3 medicines)** |  |  |
| 0 medicines | 0.58 (0.27-1.21) | 0.146 |
| 1 medicine | 1.00 (0.52-1.91) | 0.995 |
| 4-5 medicines | 1.78 (1.02-3.12) | 0.044 |
| ≥6 medicines | 1.15 (0.59-2.23) | 0.687 |
| **Urban**^a^ | 1.19 (0.78-1.81) | 0.418 |
| **Partnered**^a^ | 0.67 (0.44-1.03) | 0.418 |

Note: The variables are not mutually adjusted.

^a^Reference groups where not indicated are no LTI scheme eligibility, no private health insurance, male sex, no complex multimorbidity, not urban, and not partnered.

Abbreviations: CI, confidence interval; GMS, General Medical Services; GP, general practitioner; LTI, Long Term Illness
